# Generalizable Clinical Note Section Identification with Large Language Models

**DOI:** 10.1101/2024.02.18.24303014

**Authors:** Weipeng Zhou, Timothy A. Miller

## Abstract

**Objective:** Clinical note section identification helps locate relevant information and could be beneficial for downstream tasks such as named entity recognition. But the traditional supervised methods suffer from transferability issues. This study proposes a new framework for using large language models for section identification to overcome the limitations.

**Materials and methods:** We framed section identification as question-answering and provided the section definitions in free-text. We evaluated multiple LLMs off-the-shelf without any training. We also fine-tune our LLMs to investigate how the size and the specificity of the fine-tuning dataset impacts model performance.

**Results:** GPT4 achieved the highest F1 score of 0.77. The best open-source model (Tulu2-70b) achieved 0.64 and is on par with GPT3.5 (ChatGPT). GPT4 is also found to obtain F1 scores greater than 0.9 for 9 out of the 27 (33%) section types and greater than 0.8 for 15 out of 27 (56%) section types. For our fine-tuned models, we found they plateaued with an increasing size of the general domain dataset. We also found that adding a reasonable amount of section identification examples is beneficial.

**Discussion:** These results indicate that GPT4 is nearly production-ready for section identification, and seemingly contains both knowledge of note structure and the ability to follow complex instructions, and the best current open-source LLM is catching up.

**Conclusion:** Our study shows that LLMs are promising for generalizable clinical note section identification. They have the potential to be further improved by adding section identification examples to the fine-tuning dataset.

## 1 Introduction

Clinical notes are a rich component of electronic health records (EHR) and natural language processing (NLP) methods can help extract useful information from notes and assist clinical reasoning and knowledge finding^1,2^. Clinical notes are organized by sections and automatically identifying sections can benefit downstream tasks such as named entity recognition^3^, cohort discovery^4^ and symptom negation^5^. For instance, identifying the social history section can be helpful for the extraction of social determinants of health ^6^.

Many previous methods for section identification relied on training machine learning models on human annotated data, usually for a specific note type with a predefined list of section types.^7–9^ They achieve F1 scores as high as 0.90 in the source healthcare institution where they were annotated but usually drop significantly (as low as 0.6 F1) when transferred to another healthcare institution. Part of this could be due to the note taking differences between healthcare institutions. In addition, these models are highly restricted by the annotation schema, making it difficult to apply trained models to different note types or the same note type but written with a different structure. For example, a model trained on discharge summaries would have difficulty in applying to progress notes, because some section types (e.g., “overnight progress”), are usually not found in discharge summaries. Lastly, per-section type accuracy could be dependent on the number of annotations available^10^. For section types that are scarce, supervised learning models tend to have a lower accuracy due to insufficient annotated examples.

Recent studies that applied large language models (LLM) such as ChatGPT^11^ and GPT4^12^ to healthcare domains achieved promising results^13^. LLMs use little or no annotated data and function in a question-answering way. It answers after receiving a human written instruction which can be rewritten or revised for different scenarios. However, like other deep learning models, LLMs lack interpretability ^13^, which makes their application in high-stakes domains like medicine riskier.

## 2 Objective

In this study, we experiment with applying LLMs to section identification with two goals. First, we measure state-of-the-art LLMs performance for section identification with different accessibility (open-sourced and closed-sourced) and with varying parameter sizes. Second, we seek to better understand LLMs by analyzing their behavior on section identification, which is a comprehensive task that involves clinical knowledge understanding, verbatim copying, label assignment, and output formatting.

## 3 Method

### 3.1 Dataset

We refer to the evaluation dataset we use in this study as **discharge**^9^, which consists of discharge summaries from the Partners HealthCare and Beth Israel Deaconess Medical Center and was originally released through the i2b2 challenge. The dataset consists of 92 notes with 29 section types, which we map to 27 section types to remove redundant categories (See Supplementary Appendices). We selected 50 notes for the test set, ensuring that, when combined with the prompt, each input to the LLM is at most around 3500 tokens, so there is enough room for the model (context window at 4096 tokens) to generate responses. This helps for making comparisons between the LLMs -- although the version of GPT4 we use allows for a context window of 8k tokens, the other models are limited to 4k tokens. The rest of the notes are used for development.

To better understand factors that impact LLM on section identification, we also collected two datasets for training customized models. We used **progress**,^10,14,15^ which consists of 765 progress notes with 17 section types, which we reduce to 15 section types by mapping several categories to “Unknown.” (See Appendices Table 2) from Beth Israel Deaconess Medical Center and is released as a part of MIMIC-III. We converted the progress notes into question-answer pairs for instruction tuning as described in future sections.

We also used **ORCA**^16,17^, an instruction tuning dataset that contains question-answer pairs collected from ChatGPT/GPT4 using self-instruct. Self-instruct^18^ is the technique composing questions from existing annotated datasets and using them to query ChatGPT/GPT4 to create high-quality answers. The **ORCA** dataset used in our study contains 0.5 million question-answer pairs created using GPT4.

### 3.2 Model

#### 3.2.1 Off-the-shelf model

We evaluated GPT3.5 (ChatGPT) and GPT4^11,12^, which are the state-of-the-art closed-source models. We also evaluated state-of-the-art open-source models with 13 billion or 70 billion parameters, Llama2-13b-chat^19^, Vicuna-13b^20^ (v1.5), Llama2-70b-chat^19^, and Tulu2-70b^21^ (with direct policy optimization^22^ applied). The Llama2 models are developed by Meta^19^.

Vicuna-13b is developed by researchers in UC-Berkeley LMSYS group and is a state-of-the-art 13 billion model. Tulu2-70b is developed by Allen Institute for AI and is reported to be competitive against ChatGPT^21^.

All open-source models are tuned on the top of Llama2-13b-base or Llama2-70-base^19^. Llama2-13b-chat and Llama2-70b-chat are instruction tuned followed by reinforcement learning-based techniques for aligning with human preferences (i.e., reinforcement learning from human feedback -- RLHF). Vicuna-13b is trained on human-ChatGPT conversations users shared online^23^. Tulu2-70b is trained with a large and diverse instruction dataset followed by direct policy optimization (DPO), a technique with a similar objective as RLHF but removing the need of doing reinforcement learning^21^.

GPT3.5 has 4096 token context window size (maximum input token limit). GPT4 allows 8192 tokens but we limited it to 4096 for fair comparison. For GPT3.5 and GPT4, we are using them in a HIPAA-compliant environment provided through the Microsoft Azure cloud computing platform. Both GPT3.5 and GPT4 are accessed using the API version “2023-03- 15-preview”. The open-source models have a context window size of 4096 tokens and their querying is handled by FastChat^24^, which is an open-source LLM hosting platform deployable locally that allows querying LLM in a way similar to GPT3.5/GPT4 by internally handling each model’s special input format.

#### 3.2.2 Customized model

To better understand factors that impact LLM performance on section identification, we also train our own LLM on top of Llama2-13b-base. Similar to how Vicuna-13b is trained on Llama2-13b-base, we perform instruction tuning on Llama2-13b-base using the **ORCA** and/or **progress** dataset. We also try continued instruction tuning on top of Vicuna-13b. Due to hardware limitations, we employed LoRA^25^ for model training. LoRA is a parameter-efficient training technique that tunes only a small number of model parameters. When tuning, we used a batch size of 16, epoch size of 1, learning rate of 0.0004, rank of 16, alpha of 16, dropout of 0.05 and LoRA modules of gate, down and up, following the set up in Lee et al^26^. When training our customized model, we also vary the size and ratio of the **ORCA** and progress dataset. We train with **ORCA**, with its size varying from 25k, 50k, 100k, 250k to 500k. We also train with **progress**, with its size varying from 25, 50, 100, 250 to 500, and they are combined with an additional 25k **ORCA** examples. In preliminary experiments, we found that without any general-domain data (e.g., **ORCA**), the training was less stable and the model seemed to overfit to the **progress** dataset’s section types. We also tried continual instruction tuning on top of Vicuna-13b. We tuned it with 500 examples from **progress** and/or 25k question-answer pairs sampled from **ORCA**. Table 1 s summarizes the customized models trained in this study.

**Table 1.** Name, base model, and instruction tuning dataset size of the customized models in this study.

| Customized model name | Base model | **ORCA** sample size | **progress** sample size |
| --- | --- | --- | --- |
| Llama<sub>o</sub>-25k | Llama2-13b-base | 25k | 0 |
| Llama<sub>o</sub>-50k |  | 50k |  |
| Llama<sub>o</sub>-100k |  | 10k |  |
| Llama<sub>o</sub>-250k |  | 250k |  |
| Llama<sub>o</sub>-500k |  | 500k |  |
| Llama<sub>o</sub>-25k, p-25 |  | 25k | 25 |
| Llama<sub>o-25k, p-50</sub> |  |  | 50 |
| Llama<sub>o-25k, p-100</sub> |  |  | 100 |
| Llama<sub>o-25k, p-250</sub> |  |  | 250 |
| Llama<sub>o-25k, p-500</sub> |  |  | 500 |
| Vicuna<sub>p-500</sub> | Vicuna-13b | 0 | 500 |
| Vicuna<sub>o-25k, p-500</sub> |  | 25k |  |

### 3.3 Prompt design

The model prompt is a central part to evaluating LLMs. In this study, we designed the prompt following common practices in LLM querying as well as bypassing some known LLM limitations to make it suitable for applying to section identification. Figure 2 shows the prompt template used in this study.

**Figure 1.**
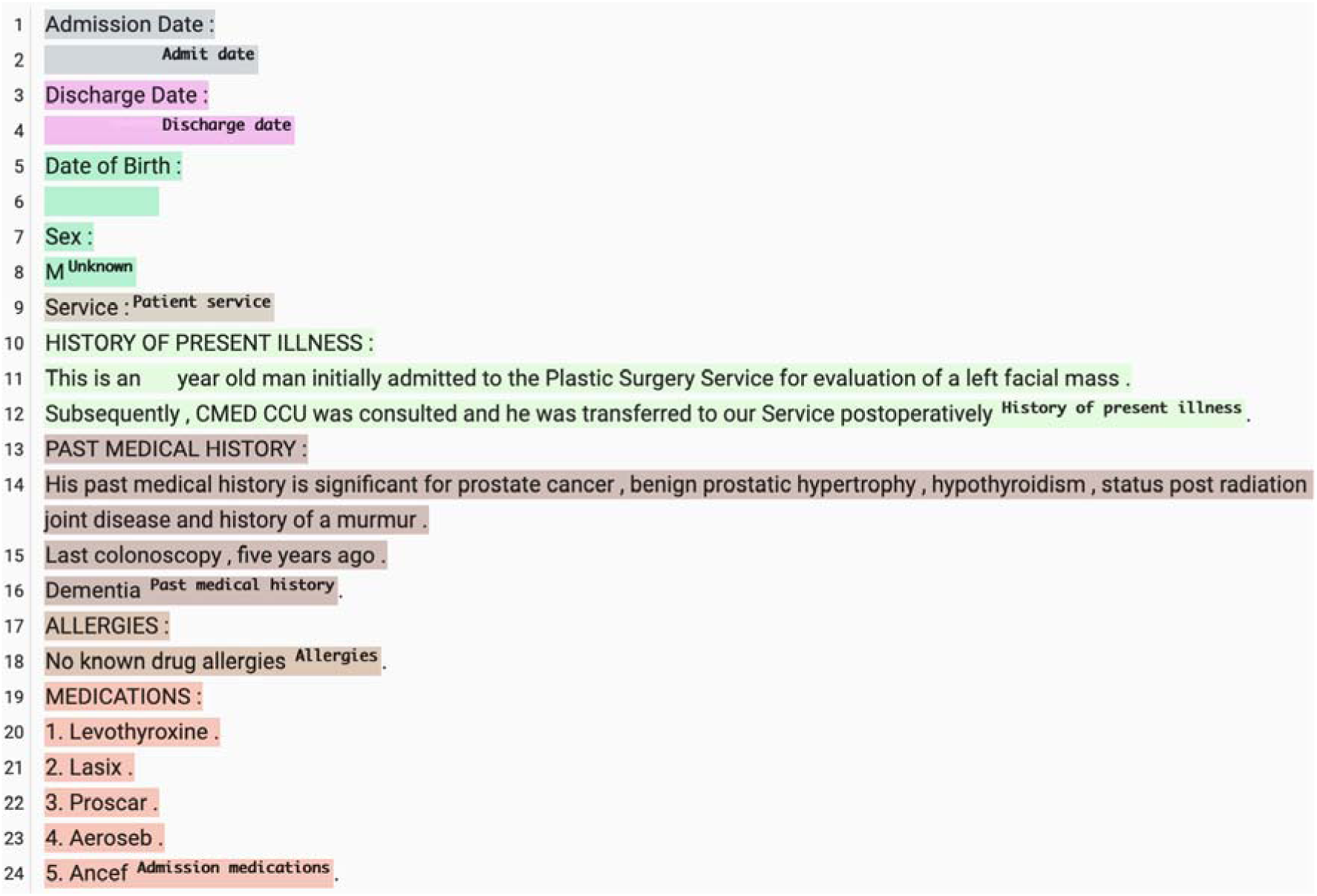
A discharge summary with sections identified by GPT4, with date and age censored.

**Figure 2.**
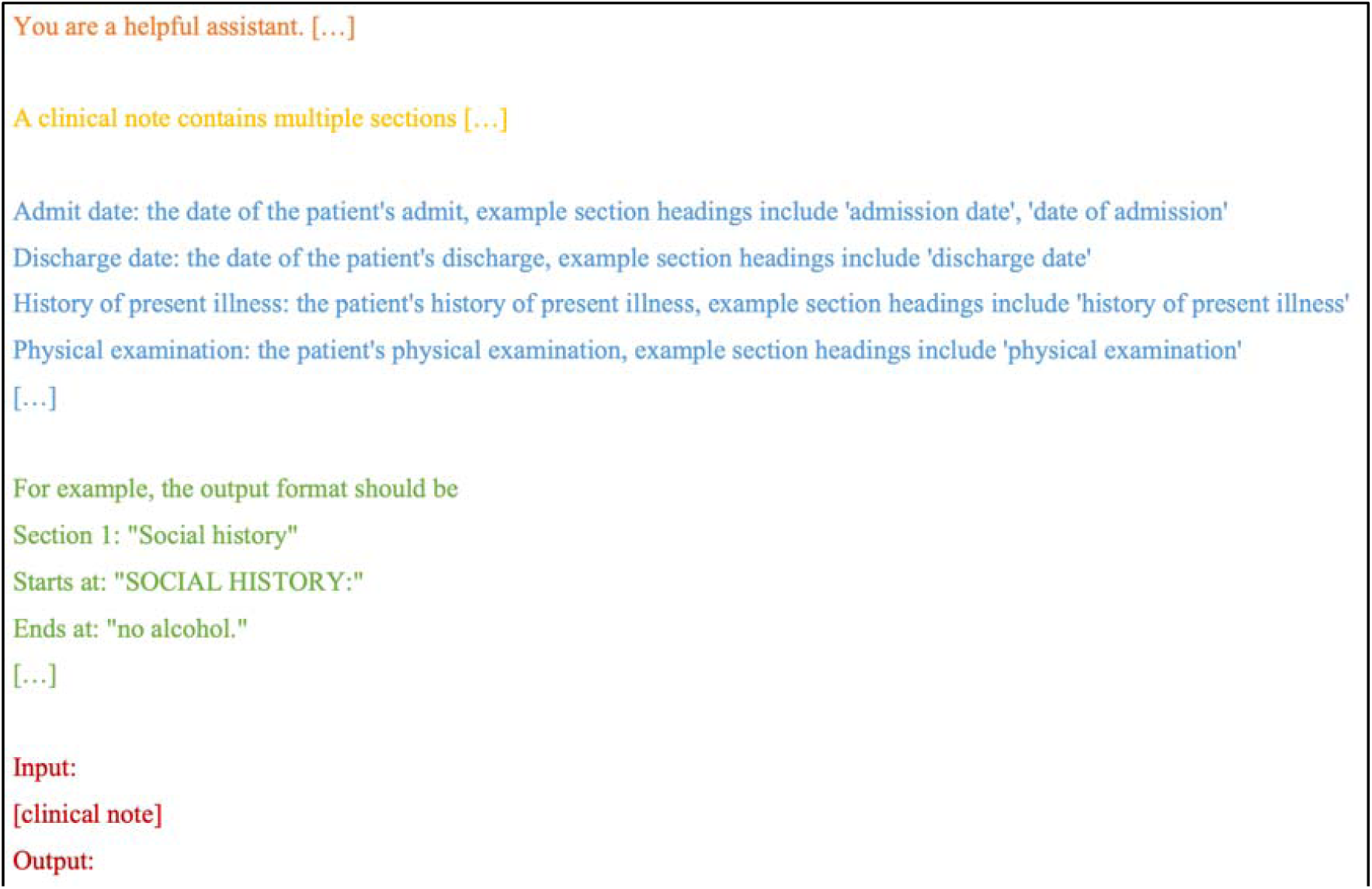
Overview of the prompt (input) provided to the large language models for section identification.

Our prompt contains four components: system message, task description, section definitions, and clinical note. An example of the prompt, as well as the example output, is provided in Supplementary Appendices. The system message is a high-level instruction to LLMs about how it should behave. In our case, the system message is provided as “You are a helpful assistant. You are an experienced clinician and you are familiar with writing and understanding clinical notes.”

Then, we provide the task description (section identification) to the LLM – “A clinical note contains multiple sections like family history, allergies and history of present illness. Given a clinical note as an input, please separate the notes into sections and output their section names. Also specify where the section starts and ends. Use section names from one of the following:”.

Following the task description, we provide section definitions. They contain the section name, the definition and example section headings. Since the **discharge** and **progress** datasets did not provide them, we derived them by inspecting the clinical notes from the non-test set. An example definition for the section type “Admit date” is “Admit date: the date of the patient’s admit, example section headings include ‘admission date’, ‘date of admission’.” We created a section type “Unknown” for section types that could cause confusion to the model, such as “Subsection”. We also modified some of the originally provided section labels to make them more understandable, such as changing “Physical” to “Physical examination.” The section mappings and definitions are included in the Appendices.

Following the section definitions, we provide two example output sections to show the LLM how to format the extracted sections. We found in preliminary experiments that they are useful for models to produce consistent and desired output formats. We instruct the LLM to format sections to begin with a section index and a section name, then the beginning text of the section, and the end text of the section. Ideally, the LLM should output the start and end character index for the section directly, but LLMs do not natively reason in character offset space and have been shown to perform poorly at generating such outputs^27,28^. Alternatively, LLMs are good at verbatim copying of text^29,30^. Following the section definitions, we provide the LLM with a clinical note followed by an “Output:” prompt which signals the LLM to output the response.

### 3.4 Output postprocessing

Even with the output format included in the prompt, our models occasionally generated unstructured outputs during development. To avoid these, we employ a mechanism that retries the generation when it does not give the desired output format at the minimum, in other words, retry when the extracted section output is not like “Section [number]: [some text] Starts at: [some text] Ends at: [some text]”.

When the format is correct, there are also content errors such as invalid section names and section start/end text. With this observation, we design an output postprocessing algorithm to convert the output to section names and character spans for evaluating against the gold annotations. Assuming that, when given a clinical note, the LLM extracts a list of sections in the predefined format, and each section contains three pieces: section name, section start text and section end text, we take the following steps.

1. Remove sections that do not have a valid section name. A valid section name is defined to be exactly matching one of the section names as described in the section description part of the prompt.
2. Remove sections that do not have a valid section start text. A valid section start text is defined to be having an exact match in the clinical note.
3. For the remaining sections, use the section start text to locate the section’s beginning character index.
4. Sort sections in the order of the section beginning character index.
5. Define a (tentative) section end character index as the next section’s beginning character index - 1.
6. Find a section’s end text in between the section’s beginning and end character index. If a match is found, use that as the section’s new end character index.

An advantage of this postprocessing method is it can still locate a section even when its section end text is incorrectly generated by the LLM, as long as the section start text is valid. Sometimes LLMs make errors in verbatim copying the section name and section start/end text. We experiment with two variations of the above algorithm to mitigate them.

*Section name ignore case match*: In step 1 when we are matching section names, instead of exact match, we use case insensitive match when finding valid section names.

*Fuzzy section start/end match*: In steps 2 and 6, when we are matching section start/end text within the clinical note, we use fuzzy match instead of exact match. The fuzzy match lists all possible sequences in the clinical note and finds the sequence most similar to the start/end text. The similarity is scored based on Levenshtein distance^31^, which measures the similarity between two sequences as the length-normalized minimum number of single-character edits between them, and it ranges from 0 to 100. We define that a match occurs if the similarity score is greater than or equal to 90.

### 3.5 Evaluation

Due to budget limitations, we evaluate GPT4 and GPT3.5 for 3 repetitions and report the mean and standard deviation. For the open-source and customized models, we repeat the evaluation 5 times. We report micro-F1 scores instead of macro-F1 because section types are not evenly distributed.

Traditional supervised learning section identification methods used BIO tagging for evaluation^9^. Limited by the computational complexity, they made sentence level prediction and evaluated based on that. However, large language models generate characters one by one and do not have this concern. It is more suitable to evaluate with a span-based approach.

Inspired by Beeferman et al.^32^, which uses an asymmetric function for evaluating section boundaries, and *nervaluate*^33^ on evaluating named entity recognition, we proposed a span-based evaluation method for section identification. This method considers both section span coverage and section label correctness.

We define an algorithm, *Evaluate (L1, L2)*, for calculating *L1*’s accuracy and match ratio against *L2*. *L1* and *L2* are both a list of sections with a span indicating the start and end character index of the section, and a section type. The algorithm is shown below. *L1* can be either the predicted sections or the target sections. *R* is a list of section match ratio (*r*) for the matched *L1* sections. *N_correct_* is the number of sections in *L1* with a correct match in *L2*. Note that *r* is calculated for every section in *L1*. This helps us understand the model’s span finding capability directly.

**Algorithm:**
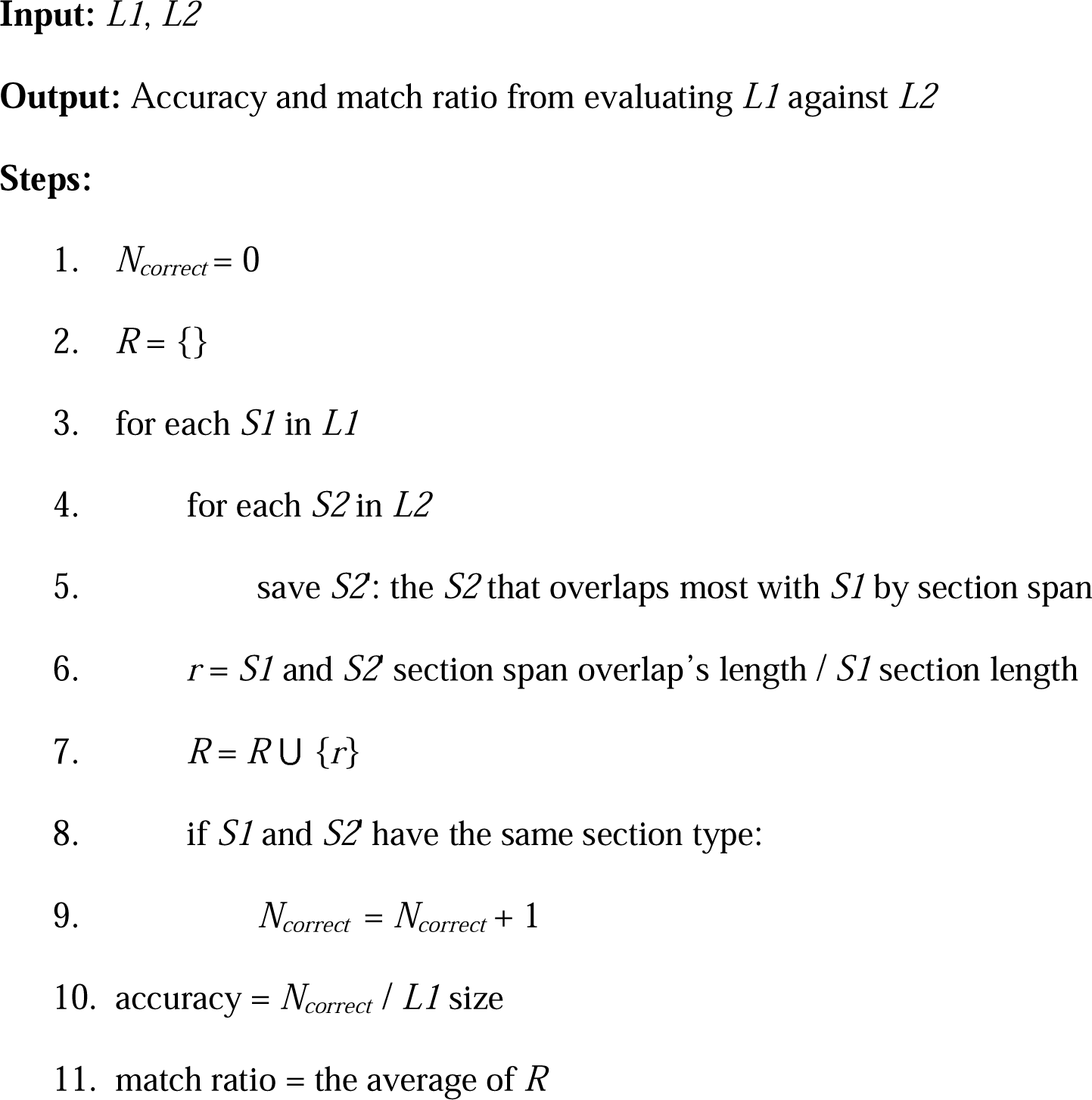
Evaluate (L1, L2)

Following the general definition of precision and recall, we define for section identification:

precision, prediction match ratio = *Evaluate (L1=predicted sections, L2=target sections)*

recall, target match ratio = Evaluate (L1=target sections, L2=predicted sections)
F1 = 2 * precision * recall / (precision + recall)
match ratio = (prediction match ratio + target match ratio) / 2

## 4 Results

### 4.1 Models

#### 4.1.1 Off-the-shelf model

Table 2 shows the results of each system on the discharge dataset. GPT4 has the highest performance (F1=0.77) while GPT-3.5 and Tulu2-70b score similarly with F1=0.64. All other models score are not competitive. In terms of the stability over runs, the GPT models are most stable, while open-source models have lower stability.

**Table 2.** The F1 and match ratio of the open-sourced and closed-sourced models, with sequential ablations of the fuzzy start/end text match and then case insensitive section name match techniques.

|  | GPT4 | GPT3.5 | Llama2-13b-chat | Vicuna-13b | Llama2-70b-chat | Tulu2-70b |
| --- | --- | --- | --- | --- | --- | --- |
| F1 (std) / Match ratio (std) | 0.77(0.003)/<br>0.91(0.003) | 0.64(0.005)/<br>0.82(0.011) | 0.39(0.014)/<br>0.59(0.014) | 0.42(0.03)/<br>0.7(0.019) | 0.46(0.012)/<br>0.69(0.012) | 0.64(0.008)/<br>0.79(0.009) |
| - Fuzzy start/end text match | 0.77(0.003)/<br>0.91(0.003) | 0.64(0.006)/<br>0.83(0.006) | 0.13(0.028)/<br>0.46(0.018) | 0.33(0.032)/<br>0.63(0.015) | 0.03(0.008)/<br>0.35(0.031) | 0.39(0.043)/<br>0.64(0.043) |
| - Case insensitive section name match | 0.77(0.003)/<br>0.91(0.003) | 0.63(0.003)/<br>0.82(0.007) | 0.02(0.009)/<br>0.26(0.039) | 0.27(0.042)/<br>0.57(0.035) | 0.02(0.007)/<br>0.33(0.051) | 0.38(0.043)/<br>0.63(0.043) |

We performed ablation studies by sequentially removing the two postprocessing enhancement techniques: *fuzzy start/end text match* and *case-insensitive section name match*. We found this had very little impact to GPT3.5 and GPT4 but enormous impact for the open-source models. Removing *fuzzy start/end text match* led to a substantial F1 score decrease (0.25) for Tulu2-70b as well as higher variations across runs, making it no longer competitive with GPT3.5. Other open-source models had similar large performance decreases. The F1 decreases to as low as 0.02 for Llama2-70b-chat.

Removing *case-insensitive section name match* further has the biggest impact on Llama-13b- chat, decreasing the F1 score from 0.12 to 0.02. Vicuna-13b’s F1 score decreases by 0.06. Little impact was observed for the 70b models.

#### 4.1.2 Customized model

Table 3 (top row) shows F1 scores for the customized LLMs instruction tuned on top of Llama2-13b-base using the **ORCA** and **progress** datasets. We found that F1 improves from 0.291 to 0.38 when changing the number of **ORCA** dataset size from 25k to 50k. The F1 remains stable with additional instances.

**Table 3.** The F1 and match ratio of customized models.

| Model | Llama<sub>o</sub>-25k | Llama<sub>o</sub>-50k | Llama<sub>o</sub>-100k | Llama<sub>o</sub>-250k | Llama<sub>o</sub>-500k |
| --- | --- | --- | --- | --- | --- |
| F1 (std) /<br>Match ratio(std) | 0.29(0.007)/<br>0.6(0.023) | 0.38(0.025)/<br>0.61(0.029) | 0.36(0.03)/<br>0.61(0.023) | 0.38(0.02)/<br>0.64(0.019) | 0.37(0.009)/<br>0.63(0.007) |
| Model | Llama<sub>o</sub>-25k, p-25 | Llama<sub>o</sub>-25k, p-50 | Llama<sub>o</sub>-25k, p-100 | Llama<sub>o</sub>-25k, p-250 | Llama<sub>o</sub>-25k, p-500 |
| F1 (std) /<br>Match ratio(std) | 0.31(0.034)/<br>0.61(0.019) | 0.36(0.041)/<br>0.62(0.026) | 0.35(0.028)/<br>0.65(0.01) | 0.32(0.023)/<br>0.63(0.015) | 0.3(0.018)/<br>0.63(0.013) |
| Model | Vicuna<sub>p</sub>-500 | Vicuna<sub>o</sub>-25k, p-500 |  |  |  |
| F1 (std) /<br>Match ratio(std) | 0.26(0.015)/<br>0.65(0.018) | 0.33(0.015)/<br>0.68(0.006) |  |  |  |

In Table 3 (bottom row), when adding domain-specific instructions to the dataset, adding 25 section identification examples from **progress** in addition to the 25k **ORCA** samples increased the F1 score slightly from 0.29 to 0.31. And increasing the number of instances to 50 and 100 further improved the F1 to around 0.35. Increasing the size of the examples to 100, 250 and 500 resulted in a F1 drop until 0.3.

In experiments where we tried to continue instruction tune on top of Vicuna-13b, a model already instruction tuned on top of Llama2-13b-base, we found that both continuous instruction tuning methods (Vicuna_p-500_ and Vicuna_o-25k, p-500_) resulted in a performance drop.

## 5 Discussion

### 5.1 Analysis of GPT4 results

We take an in-depth look into GPT4’s behavior to gain more insight about LLMs in section identification. We randomly selected one run for analysis. Table 4 shows the F1 scores by section types. There are 27 section types in total and we observed that 9 (33%) have an F1 score greater than 0.9, and 15 (56%) have an F1 score greater than 0.8. These are often common and important section types such as “Social history” and “Past medical history.”

**Table 4.** The detailed performance values of one randomly selected GPT4 run after breaking down into section types, sorted by F1 scores.

| Section type | Precision | Recall | F1 | Prediction section count | Prediction match ratio | Target section count | Target match ratio |
| --- | --- | --- | --- | --- | --- | --- | --- |
| Social history | 1 | 1 | 1 | 15 | 1 | 15 | 0.98 |
| Family history | 1 | 1 | 1 | 8 | 1 | 8 | 0.98 |
| Gynecologic history | 1 | 1 | 1 | 3 | 1 | 3 | 0.98 |
| Admit date | 0.98 | 0.98 | 0.98 | 50 | 1 | 50 | 0.56 |
| Past medical history | 0.96 | 0.96 | 0.96 | 27 | 1 | 28 | 0.91 |
| Other diagnosis | 0.95 | 0.95 | 0.95 | 20 | 0.98 | 20 | 0.74 |
| Past surgical history | 1 | 0.89 | 0.94 | 8 | 1 | 9 | 0.98 |
| Discharge condition | 0.89 | 0.96 | 0.92 | 27 | 1 | 25 | 0.96 |
| Allergies | 0.92 | 0.92 | 0.92 | 24 | 1 | 24 | 0.74 |
| Discharge date | 0.98 | 0.81 | 0.89 | 50 | 0.97 | 57 | 0.38 |
| Discharge medications | 0.87 | 0.89 | 0.88 | 38 | 1 | 36 | 0.75 |
| History of present illness | 0.86 | 0.86 | 0.86 | 29 | 1 | 29 | 0.99 |
| Admission medications | 0.9 | 0.79 | 0.84 | 21 | 1 | 24 | 0.98 |
| Patient procedures | 0.85 | 0.84 | 0.84 | 33 | 0.95 | 31 | 0.86 |
| Hospital course | 0.7 | 1 | 0.83 | 47 | 1 | 37 | 0.95 |
| Physical examination | 0.64 | 0.96 | 0.77 | 25 | 0.91 | 25 | 0.93 |
| Lab studies | 1 | 0.58 | 0.74 | 16 | 1 | 24 | 0.74 |
| Discharge diagnosis | 0.87 | 0.64 | 0.73 | 23 | 1 | 33 | 0.86 |
| Admission diagnosis | 0.61 | 0.83 | 0.7 | 31 | 1 | 23 | 0.81 |
| Patient comments | 0.8 | 0.6 | 0.69 | 10 | 0.99 | 15 | 0.78 |
| Reason for admission | 0.75 | 0.62 | 0.68 | 12 | 0.89 | 13 | 0.98 |
| Patient service | 0.85 | 0.56 | 0.68 | 13 | 1 | 16 | 0.73 |
| Attending physician | 0.44 | 0.79 | 0.57 | 34 | 0.98 | 19 | 0.51 |
| Follow-up | 0.63 | 0.49 | 0.55 | 32 | 0.99 | 43 | 0.58 |
| Discharge instructions | 0.56 | 0.45 | 0.5 | 25 | 0.98 | 29 | 0.71 |
| Admitting physician | 0.05 | 1 | 0.1 | 19 | 1 | 1 | 0.25 |
| Unknown | 0.25 | 0 | 0 | 4 | 1 | 61 | 0.62 |

Nevertheless, we found GPT4 still scores below or equal to 0.1 F1 for two section types (7%). The lowest performing section type for GPT4 is “Unknown” with F1 being 0. There are four predictions and 61 gold annotations. When matching the predictions against gold annotations, only one section was matched with correct section name. When matching the gold annotations against predictions, no section was matched with correct section names. This might be due to the different interpretations of “Unknown.” The gold annotations tend to make the header of the note as “Unknown” sections, while GPT4 tends to label it for sections that have contents that it could not understand, following what the prompt defines. Another low-performance section type is “Admitting physician” (F1=0.1). GPT4 made 19 predictions for “Admitting physician” and the gold annotations only have one instance annotated. We find that GPT4 tended to consider the sections that start with “Dictated by”/“Dictating for” and contains a physician name as the “Admitting physician” section. Those sections are also usually ignored in the gold annotations, resulting in the F1 prediction and gold annotation mismatch that we observed.

Additional qualitative assessment shows that GPT4 might be better than what the F1 score suggests. Sometimes GPT4’s misclassification comes from it interpreting section definition differently from the gold annotation. The gold annotation tends to annotate the whole note, and GPT4 tends to only annotate the parts that it is sure about. For example, on the left of Figure 4 is the gold annotation, and it annotates two “Discharge date” sections, in contrast to one section from GPT4 but essentially the same information. Both sections from the gold annotations are wider than what’s annotated by GPT4, but it is difficult to say that GPT4 is wrong, and it depends on the use case.

**Figure 3.**
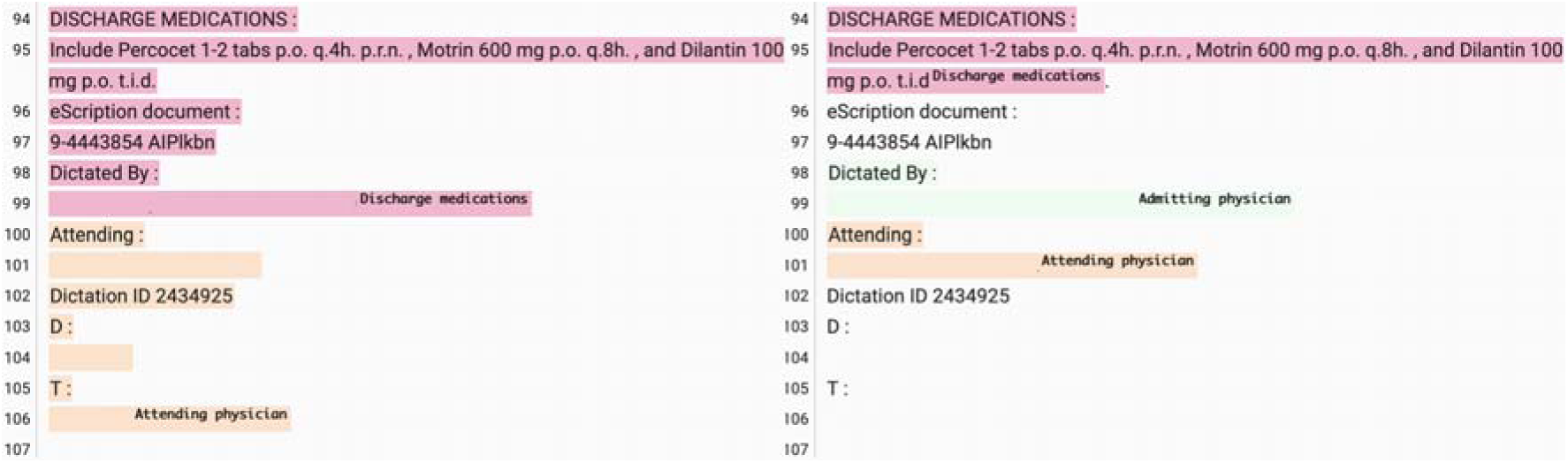
An example of “Admitting physician” annotation difference between the human annotator (left) and GPT4 (right), censoring name and date.

**Figure 4.**
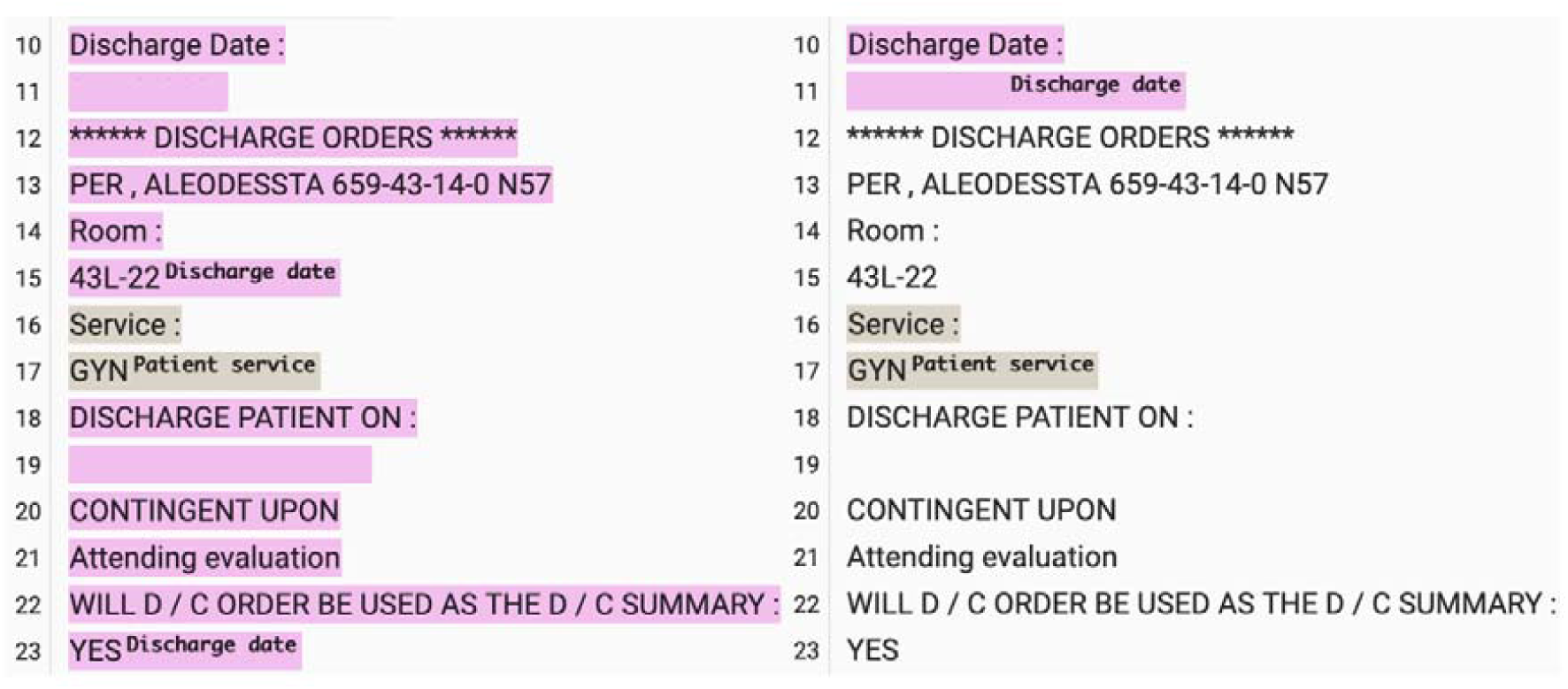
Comparing human (left) and GPT4 (right) annotated clinical notes on discharge dates, with date censored.

When GPT4 does not match the gold annotations, it is sometimes an annotation error. In the below example, the gold annotation misannotated the “Discharge medication” as “Admission medication”, while GPT4 annotated it correctly.

Traditional supervised learning models’ accuracy for each section type tends to be higher if more annotations are available, and lower when fewer annotations are available^10^. For GPT4, we found that the model does not have a clear association with a section’s annotation counts (Figure 6).

**Figure 5.**
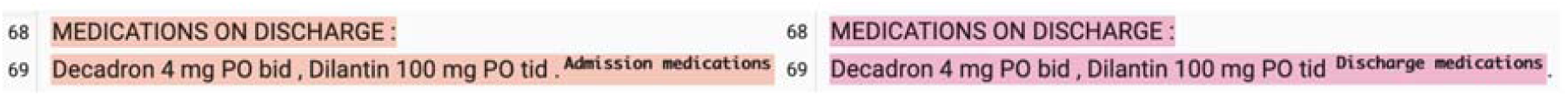
An example of the “Discharge medication” section that is annotated wrongly by human but correctly by GPT4.

**Figure 6.**
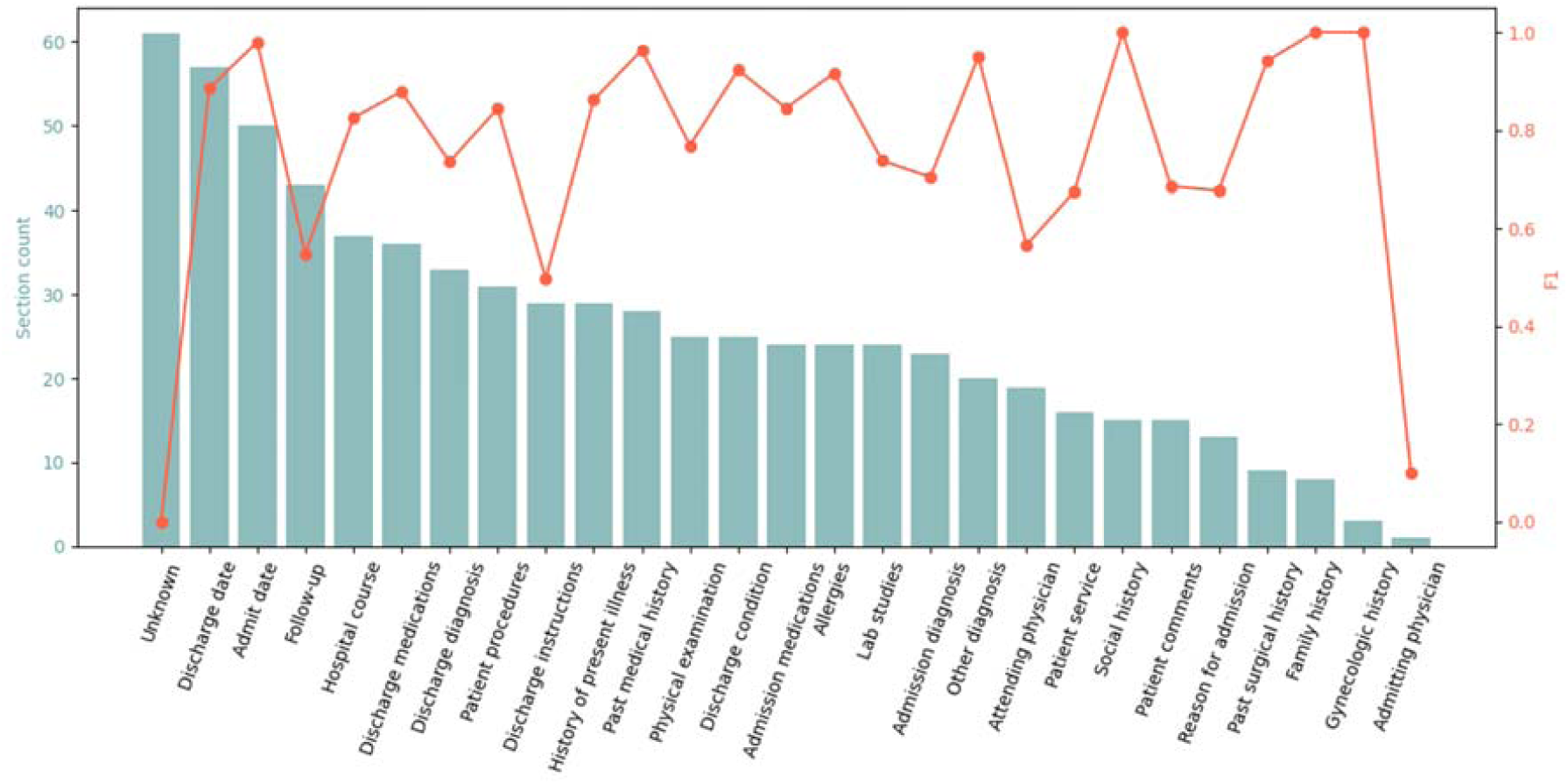
An illustration of non-dependent relationship between section counts (left y-axis) and F1 scores (right y-axis) for GPT4’s predictions. The x-axis is the section types.

### 5.2 Open-source models are closing the performance gap

All open-source models experienced a large performance drop when removing the fuzzy *section start/end match* in postprocessing; however, it is worth noticing that Vicuna-13b has the least performance drop across the open-source models. Vicuna-13b differs from the other three in that it only performs instruction tuning, without following RLHF or DPO. This might indicate that a second tuning stage based on human preference has the potential of reducing a model’s verbatim copying ability. An explanation might be the so-called “Alignment tax^34^”, where fine-tuned LLMs lose performance on language modeling tasks due to aligning with human preferences for better performance in chat applications. When removing the *case-insensitive section name match* further so that the models are evaluated without any postprocessing, both 13b models’ performance continue to decrease, but not for the 70b models. When comparing Tulu2-70b to the 13b models, this might indicate that larger models are better at copying the provided section names while keeping the original upper and lower cases.

Since the release of ChatGPT in the end of 2023, the LLM research community has been working towards reproducing it with open-source efforts, and the competitive performance of Tulu-2-70b against GPT3.5 in section identification is encouraging. However, we do notice that this is achieved with noticeable postprocessing efforts. Both accuracy and stability of Tulu-2-70b decreased without postprocessing while GPT3.5 and GPT4 still perform consistently.

### 5.3 Comparing between open-source models

When comparing between the open-source models, we found the two Llama-chat models are consistently underperforming from their same size equivalence. There are many differences in model development that could explain these results, but one important factor to keep in mind is that the Llama models were released earlier and the later-released models may have learned from their predecessors^19–21^.

### 5.4 Efficient training of customized models

When training the customized LLMs with an increasing size of general domain dataset (**ORCA**), we found the resulted F1 score improves at the beginning, but the gain becomes diminishing afterwards. Alternatively, we experimented with training the LLM with a small amount of section identification examples, and found they are effective in improving the F1 score. In the future, when people are trying to train LLMs for section identification, a cost-effective approach might be to train the model with a modest amount of general domain dataset and some section identification examples combined. It is also worth noticing that one should be careful about balancing the ratio of general domain dataset and section identification examples, as our experiments found that adding too many section identification examples can lead to a decreased model performance, likely due to overfitting.

Applying continued instruction tuning on top of already instruction-tuned LLM (Vicuna-13b) led to decreased model performance. This indicates that this approach could be sensitive to overfitting despite the strong initialization point. The performance drop of Vicuna_o-25k, p-500_, however, is lower than that of Vicuna_p-500_. This might be because Vicuna_o-25k, p-500_ is trained with an additional 25k general domain instances, alleviating the overfitting.

### 5.5 Limitations

Our study shows the effectiveness of LLM in section identification; however, due to hardware limitation, we only experimented with applying off-the-shelf models on discharge summaries, or training 13b models with parameter efficient training techniques (LoRA^25^) on progress notes. To better understand LLM in section identification, future studies can consider extending the methodology to other types of clinical notes such as radiology reports, as well as training larger models and using full fine-tuning. Second, our study limits models’ input token size to 4096 to enable straightforward comparisons between LLMs. GPT4 has a longer input limit and more expensive versions of GPT3.5 supports longer input. There are also open-sourced models that allow for longer inputs^35–37^. Understanding LLM’s section identification performance for longer notes, or multiple notes in concatenation, could be a meaningful area.

## 6. Conclusions

We evaluated the use of LLM for section identification under a highly transferrable framework. Our experiments show that GPT4 performed the best and achieved scores of 0.9 F1 or better for one third of the section types on a discharge summary dataset. Experiments suggest that the customized LLMs plateaued with an increasing number of the general domain instructions, and adding a reasonable amount of section identification examples is effective for improving model’s performance.

## 7. Author contributions

W.Z. and T.M. designed the study. W.Z. carried out all of the experiments in the study under the guidance of T.M. and created the manuscript.

## 10. Data availability

The discharge dataset notes are available at https://portal.dbmi.hms.harvard.edu/projects/n2c2-nlp/, and the section annotations can be found at https://github.com/2533245542/SectionChunker_i2b2_2010_data. The progress dataset is available at https://physionet.org/content/task-1-3-soap-note-tag/1.0.0/. The ORCA dataset is available at https://huggingface.co/datasets/Open-Orca/OpenOrca.

